# The Evolution of Young People’s Mental Health during COVID-19: Evidence from four Low-and-Middle-Income-Countries

**DOI:** 10.1101/2021.06.28.21259620

**Authors:** Catherine Porter, Annina Hittmeyer, Marta Favara, Douglas Scott, Alan Sánchez

## Abstract

**Background:** Though COVID-19 presents less risk to young people of serious morbidity or mortality, the resulting economic crisis has impacted their livelihoods. There is relatively little evidence on young people’s mental health in Low-and-Middle-Income-Countries (LMICs) as the pandemic has progressed.

**Methods:** Two consecutive phone-surveys (August/October and November/December 2020) in Ethiopia, India, Peru and Vietnam interviewed around 9,000 participants of a 20-year cohort study who grew up in poverty (now aged 19 and 26). We investigate how young people’s mental health has evolved in the four countries during the pandemic. Rates of (at least mild) anxiety (depression) measured by GAD-7 (PHQ-8) were compared across countries; between males/females, and food secure/food insecure households.

**Results:** Overall, rates of at least mild anxiety (depression) significantly decreased in all countries but Ethiopia as infection rates fell. However, young people in food insecure households report high rates of anxiety and depression and have not shown consistent improvements. Food insecure households are poorer, and have significantly more children (p<0.05) except in Ethiopia.

**Conclusions:** Food insecurity has increased during the COVID-19 pandemic and is negatively associated with young people’s mental health. Urgent support is needed for the most vulnerable.

## Introduction

The COVID-19 outbreak has impacted physical and mental health across the globe. Whilst young people are relatively less vulnerable to the health effects of the virus, their education, work and social lives have been interrupted,^1^ and their futures are uncertain. Pre-pandemic research shows that 75% of mental health conditions develop by early adulthood,^2^ and that the poorest are more likely to suffer from anxiety and depressive disorders.^3^ During the pandemic, young people, especially women, experienced the greatest deterioration in mental health. ^4,5^ Tracking mental health of young people during the pandemic is critical to prevention, especially in Low-and-Middle-Income-Countries (LMICs), where young people represent one fifth of the total population, and mental health support is limited. This study outlines how young people’s mental health has evolved in four LMICs during the pandemic, provides preliminary descriptive evidence of the association between food insecurity and mental health, and briefly describes the characteristics of the food insecure.

## Methods

We analyse longitudinal data from two phone surveys conducted in Ethiopia, India (Andhra Pradesh and Telangana), Peru, and Vietnam during August-October and November-December 2020.^6^ Our sample contains participants aged 18-19 and 25-26, including 4,702 males (51%) and 4,493 females (49%) from the Young Lives study,^7^ a poverty-focused cohort study, established in 2002. 10,599 individuals were contacted, 9,195 participated in both surveys, non-response to mental health questions was rare. The main cause of attrition was the communication shutdown in Ethiopia’s Tigray region from November 2020.

Symptoms of anxiety and depression are measured in both calls using the Generalized Anxiety Disorder-7 (GAD-7) scale and the Patient Health Questionnaire depression scale-8 (PHQ-8) with a cut-off of ≥5 representing (at least) mild anxiety^8^ (depression).^9^ If any GAD-7 (PHQ-8) items were unanswered, the whole score was omitted. We define food insecurity as the household having reported running out of food at least once since the beginning of the pandemic. We characterise food insecure households according to their dwelling attributes, location, and the number of children aged 17 or under. The Home Environment for Protection (HEP)-dwelling-attribute index measures how effectively a person’s home provides protection against the coronavirus and limits the need to travel outdoors, and is thus connected with housing quality and economic status.^10, 11^ We split the index into a binary variable indicating below or above median (=low/high HEP-group) outcomes. We use *t*-tests for the difference in mean outcomes between calls, within groups, and consider a 2-sided *p*-value <0.05 as significant. Analysis is performed using Stata 14.2.

## Results

After reaching relatively high levels in mid-2020, mental health improved significantly between mid- and end-2020, in all countries except Ethiopia. Peru still had the highest rates of anxiety/depression (32% (95% CI, 29.42-33.59)/27% (95% CI, 24.65-28.62)); Vietnam the lowest (5% (95% CI, 3.95-5.63)/6% (95% CI, 5.29-7.19)) (Table I). Except Ethiopia (and for depression in India), both males and females show significant improvements. Females still report comparatively higher rates everywhere except Ethiopia, but the gender gap is closing. The proportion of households running out of food during the pandemic ranges between 7% (95% CI, 6.03-8.08) in Vietnam and 28% (95% CI, 25.78-29.68) in Ethiopia. Individuals living in food insecure households had higher rates of anxiety and depression than other groups (all countries). Mental health among the food insecure had also not necessarily improved, while other groups reported either significant improvements, or no significant increase (Ethiopia). In Peru, 46% (95% CI, 41.53-50.74) (39% (95% CI, 34.35-43.36)) of the food insecure report anxiety (depression) symptoms. Even in Vietnam, the rates of anxiety/depression (17% (95% CI, 11.07-22.26) /24% (95% CI, 17.72-30.56)) are four times the average.

**Table I:** Comparison of rates of at least mild anxiety and depression across survey periods.

|  |  | No. (%<sup>b</sup>, 95% CI) | No. at least mild anxiety (%<sup>c</sup>; 95% CI) |  | P-value | No. at least mild depression (%<sup>c</sup>; 95% CI) |  | P-value |
| --- | --- | --- | --- | --- | --- | --- | --- | --- |
|  |  |  | Aug-Oct | Nov-Dec |  | Aug-Oct | Nov-Dec |  |
| Ethiopia | Total | 2065 | 405 (19.61; 17.9-21.33) | 465 (22.52; 20.72-24.32) | 0.004 | 364 (17.63; 15.98-19.27) | 400 (19.37; 17.66-21.08) | 0.072 |
|  | Male | 1095 (53.03; 50.85-55.20) | 208 (19; 16.67-21.32) | 258 (23.56; 21.04-26.08) | 0.001 | 185 (16.89; 14.67-19.12) | 208 (19; 16.67-21.32) | 0.110 |
|  | Female | 970 (46.97; 44.80-49.15) | 197 (20.31; 17.77-22.85) | 207 (21.34; 18.76-23.92) | 0.488 | 179 (18.45; 16.01-20.9) | 192 (19.79; 17.28-22.31) | 0.350 |
|  | Not FI<sup>a</sup> | 1493 (72.3; 70.32-74.22) | 240 (16.08; 14.21-17.94) | 248 (16.61; 14.72-18.5) | 0.629 | 221 (14.8; 13-16.61) | 217 (14.53; 12.74-16.32) | 0.806 |
|  | FI | 572 (27.7; 25.78-29.68) | 165 (28.85; 25.12-32.57) | 217 (37.94; 33.95-41.93) | 0.000 | 143 (25; 21.44-28.56) | 183 (31.99; 28.16-35.83) | 0.001 |
| India | Total | 2735 | 309 (11.3; 10.11-12.49) | 228 (8.34; 7.3-9.37) | 0.000 | 274 (10.02; 8.89-11.14) | 239 (8.74; 7.68-9.80) | 0.054 |
|  | Male | 1421 (51.96; 50.06-53.84) | 134 (9.43; 7.91-10.95) | 95 (6.69; 5.39-7.99) | 0.002 | 138 (9.710; 8.17-11.25) | 109 (7.67; 6.29-9.06) | 0.019 |
|  | Female | 1314 (48.04; 46.16-49.94) | 175 (13.32; 11.48-15.16) | 133 (10.12; 8.49-11.75) | 0.002 | 136 (10.35; 8.70-12) | 130 (9.890; 8.27-11.51) | 0.651 |
|  | Not FI | 2216 (81.02; 79.50-82.48) | 229 (10.33; 9.07-11.6) | 163 (7.36; 6.270-8.44) | 0.000 | 210 (9.48; 8.26-10.7) | 181 (8.17; 7.03-9.31) | 0.073 |
|  | FI | 519 (18.98; 17.52-20.50) | 80 (15.41; 12.3-18.53) | 65 (12.52; 9.67-15.38) | 0.128 | 64 (12.33; 9.49-15.17) | 58 (11.18; 8.460-13.89) | 0.467 |
| Peru | Total | 1911 | 775 (40.55; 38.35-42.76) | 602 (31.5; 29.42-33.59) | 0.000 | 599 (31.34; 29.26-33.43) | 509 (26.64; 24.65-28.62) | 0.000 |
|  | Male | 963 (50.39; 48.13-52.66) | 319 (33.13; 30.15-36.1) | 222 (23.05; 20.39-25.72) | 0.000 | 260 (27; 24.19-29.81) | 204 (21.18; 18.6-23.77) | 0.000 |
|  | Female | 948 (49.61; 47.34-51.87) | 456 (48.1; 44.91-51.29) | 380 (40.08; 36.96-43.21) | 0.000 | 339 (35.76; 32.7-38.82) | 305 (32.17; 29.19-35.15) | 0.024 |
|  | Not FI | 1458 (76.3; 74.32-78.19) | 524 (35.94; 33.47-38.41) | 393 (26.95; 24.67-29.24) | 0.000 | 399 (27.37; 25.08-29.66) | 333 (22.84; 20.68-25) | 0.000 |
|  | FI | 453 (23.7; 21.81-25.68) | 251 (55.41; 50.81-60) | 209 (46.14; 41.53-50.74) | 0.000 | 200 (44.15; 39.56-48.74) | 176 (38.85; 34.35-43.36) | 0.034 |
| Vietnam | Total | 2484 | 229 (9.220; 8.08-10.36) | 119 (4.79; 3.95-5.63) | 0.000 | 233 (9.380; 8.23-10.53) | 155 (6.24; 5.29-7.19) | 0.000 |
|  | Male | 1223 (49.24; 47.25-51.22) | 94 (7.69; 6.19-9.18) | 55 (4.5; 3.33-5.66) | 0.000 | 93 (7.600; 6.12-9.09) | 60 (4.91; 3.69-6.12) | 0.001 |
|  | Female | 1261 (50.76; 48.78-52.75) | 135 (10.71; 9-12.41) | 64 (5.08; 3.86-6.29) | 0.000 | 140 (11.1; 9.370-12.84) | 95 (7.53; 6.07-8.99) | 0.000 |
|  | Not FI | 2310 (93.00; 91.92-93.97) | 178 (7.71; 6.62-8.79) | 90 (3.9; 3.11-4.69) | 0.000 | 185 (8.01; 6.9-9.12) | 113 (4.89; 4.01-5.77) | 0.000 |
|  | FI | 174 (7.00; 6.03-8.08) | 51 (29.31; 22.48-36.14) | 29 (16.67; 11.07-22.26) | 0.001 | 48 (27.59; 20.88-34.29) | 42 (24.14; 17.72-30.56) | 0.305 |

### Who are the food insecure?

Across countries, the food insecure are poorer and less able to protect themselves from COVID-19. In all countries except Ethiopia households with children were more vulnerable; in India, rural households (Table II).

**Table II:** Who are the food insecure?

|  |  | No. (%<sup>b</sup>, 95% CI) | Household ran out of food between the outbreak of the COVID-19 pandemic and Nov/Dec (%<sup>c</sup>; 95% CI) |  | p-value |
| --- | --- | --- | --- | --- | --- |
|  |  |  | Did not run out of food | Ran out of food |  |
| Ethiopia | Urban<sup>a</sup> | 956 (46.3; 44.13-48.47) | 698 (46.75; 44.22-49.28) | 258 (45.1; 41.02-49.19) | 0.502 |
|  | High-HEP | 1132 (54.82; 52.64-56.98) | 860 (57.6; 55.09-60.11) | 272 (47.55; 43.45-51.65) | 0.000 |
|  | Children | 1.64 (1.46) | (1.67; 1.6-1.74) | (1.56; 1.44-1.68) | 0.120 |
| India | Urban | 786 (28.74; 27.05-30.48) | 688 (31.05; 29.12-32.97) | 98 (18.88; 15.51-22.25) | 0.000 |
|  | High-HEP | 1694 (61.94; 60.09-63.76) | 1458 (65.79; 63.82-67.77) | 236 (45.47; 41.18-49.76) | 0.000 |
|  | Children | 0.82 (1.06) | (0.78; .74-.82) | (1.01; 0.9-1.11) | 0.000 |
| Peru | Urban | 1577 (82.52; 80.74-84.20) | 1214 (83.26; 81.35-85.18) | 363 (80.13; 76.45-83.81) | 0.125 |
|  | High-HEP | 1834 (95.97; 95.00-96.81) | 1416 (97.12; 96.26-97.98) | 418 (92.27; 89.81-94.74) | 0.000 |
|  | Children | 1.34 (1.32) | (1.3; 1.24-1.37) | (1.47; 1.33-1.6) | 0.020 |
| Vietnam | Urban | 1289 (51.89; 49.91-53.87) | 1208 (52.29; 50.26-54.33) | 81 (46.55; 39.12-53.99) | 0.144 |
|  | High-HEP | 2142 (86.23; 84.81-87.57) | 2023 (87.58; 86.23-88.92) | 119 (68.39; 61.46-75.32) | 0.000 |
|  | Children | 0.65 (0.97) | (0.63; 0.59-0.67) | (0.96; 0.78-1.14) | 0.000 |

Notes: <sup>a</sup>Urban= Participants living in an urban location (as opposed to rural), High-HEP= Participants fall into a high HEP-group, Children=Number of children in the household aged 17 or under.
<sup>b</sup> Mean in % in reference to the total sample. For continuous variables (number of children aged 17 or younger) mean is not in %. Number in brackets is standard deviation.
<sup>c</sup> Mean in % in reference to the respective subgroup sample. For continuous variables (number of children aged 17 or younger) mean is not in %. No.=Number. Number is omitted for continuous variables (number of children aged 17 or younger).
95% CI= 95% confidence interval
p-values represent significance of t-test of equality between households who did not run
out of food at least once between the outbreak of the COVID-19 pandemic and Nov/Dec (second phone survey) and those who did across basic household characteristics.

## Discussion

Rates of anxiety/depressive symptoms are highest in Peru and lowest in Vietnam, mirroring COVID-19 mortality rates (based on COVID-19 cases/deaths per capita).^12^ Overall, rates of anxiety and depressive symptoms in young people have significantly decreased in all countries except Ethiopia. Improvements in mental health are potentially associated with the decline in infection/mortality rates^12^ and easing of lockdown measures. Mental health worsened during the pandemic, according to studies in Germany^13^ and Poland^14^, though improved in the UK from April to June 2020.^15^

Women reported worse mental health, in all countries but Ethiopia. We note that the Ethiopian National Health Survey shows no significant differences between males and females.^16^ We find the gender gap closed as the pandemic progressed. Substantial pre-pandemic evidence indicates that women report worse mental health than men ^17, 18^ and one study observes a widening gender gap early in the pandemic in Poland^14^, mirroring evidence on gender disparities in mental health before and after natural disasters.^19^

Building on pre-pandemic research,^20^ we highlight the association between food shortages and continuing high rates of poor mental health. Young Lives research shows food insecurity increased during the pandemic, except in Vietnam^1^ and that pandemic-related stressors worsened mental health in mid-2020^21^. A 149-country pre-pandemic study^20^ found that food insecurity is associated with poorer mental health and identifies channels through which food insecurity affects mental health: i) feelings of powerlessness, sadness, frustration ii) uncertainty regarding acquiring sufficient food iii) feelings of guilt/shame from acquiring food in socially unacceptable ways.^20^

The main limitation of this study is the lack of causal inference, given the potential bidirectional relationship between food insecurity and mental health. Further, we do not capture the multidimensionality of food insecurity.

The world’s food insecure are severely, and continuously affected by the COVID-19 pandemic. Cash transfer programs (CTPs) should be expanded, and conditionality reduced/waived in LMICs to address immediate needs around food insecurity and help break the vicious cycle between the onset/persistence of mental health disorders and poverty. Further, CTPs could distribute accurate, non-stigmatised information about COVID-19 and access to mental health services using their existing communication tools.^22^ More research is needed into factors which contribute to mental health resilience during the pandemic and non-COVID-19 stressors in Ethiopia.

## Data Availability

The entire individual participant data collected during the phone survey and previous in-person rounds, after de-identification, is available including data dictionaries. Furthermore the questionnaire, attrition reports and the field work manual are available at https://www.younglives.org.uk/ . The data is available, with no end date to anyone who wishes to access the data for any purpose, via the UK Data Archive (study number 8678, DOI: 10.5255/UKDA-SN-8678-3).

https://doi.org/10.5255/UKDA-SN-8678-3

## Article Information

### Author Contributions

CP, AH, and MF conceived the study. CP, MF, DS, and AS designed the study. MF and AS led piloting of instruments and data collection. AH and CP did the statistical analyses. CP and AH wrote the first draft of the Article. MF, DS, and AS provided comments and input to the several drafts of the manuscript. MF, DS, and AS verified the underlying data. All authors critically reviewed the manuscript and approved the final draft.

### Data availability

The entire individual participant data collected during the phone survey and previous in-person rounds, after de-identification, is available including data dictionaries. Furthermore the questionnaire, attrition reports and the field work manual are available at https://www.younglives.org.uk/. The data is available, with no end date to anyone who wishes to access the data for any purpose, via the UK Data Archive (study number 8678, DOI: <u>10.5255/UKDA-SN-8678-3</u>).

### Ethics committee approval

The survey was approved by the institutional research ethics committees at the University of Oxford (UK, Ref No: CUREC 1A/ ODID CIA-20-034, 15.03.2020), the University of Addis Ababa (Ethiopia), the Centre for Economic and Social Studies in Hyderabad (India), the Instituto de Investigación Nutricional (Peru) and the Hanoi University of Public Health (Vietnam). Participants were asked for their verbal informed consent before the study commenced and were assured of confidentiality. A consultation guide was provided to all participants with resources for support in issues raised by the questionnaire, including mental health.

